# Three-dose mRNA-1273 vaccination schedule: sufficient antibody response in majority of immunocompromised hematology patients

**DOI:** 10.1101/2022.04.08.22273602

**Authors:** Sabine Haggenburg, Quincy Hofsink, Birgit I. Lissenberg-Witte, Annoek E.C. Broers, Jaap A. van Doesum, Rob S. van Binnendijk, Gerco den Hartog, Michel S. Bhoekhan, Nienke J.E. Haverkate, Judith A. Burger, Joey H. Bouhuijs, Gaby P. Smits, Dorine Wouters, Ester M.M. van Leeuwen, Hetty J. Bontkes, Neeltje A. Kootstra, Sonja Zweegman, Arnon P. Kater, Mirjam H.M. Heemskerk, Kaz Groen, Tom van Meerten, Pim G.N.J. Mutsaers, Tim Beaumont, Marit J. van Gils, Abraham Goorhuis, Caroline E. Rutten, Mette D. Hazenberg, Inger S. Nijhof, the COBRA KAI study team

**Affiliations:** Department of Hematology, Amsterdam UMC location University of Amsterdam, Amsterdam, The Netherlands; Amsterdam institute for Infection and Immunity, Amsterdam UMC, Amsterdam, The Netherlands; Department of Epidemiology and Data Science, Amsterdam UMC location Vrije Universiteit, Amsterdam, The Netherlands; Department of Hematology, Erasmus MC Cancer Institute, Rotterdam, The Netherlands; Department of Hematology, University Medical Center Groningen, University of Groningen, Groningen, The Netherlands; Centre for Immunology of Infectious Diseases and Vaccines, National Institute for Public Health and the Environment, Bilthoven, The Netherlands; Department of Experimental Immunology, Amsterdam UMC location University of Amsterdam, Amsterdam, The Netherlands; Department of Medical Microbiology and Infection Prevention, Amsterdam UMC, University of Amsterdam, Amsterdam, The Netherlands; Central Diagnostic Laboratory, Amsterdam UMC, Amsterdam, The Netherlands; Laboratory Medical Immunology, Amsterdam UMC, Amsterdam, The Netherlands; Department of Hematology, Amsterdam UMC location Vrije Universiteit, Amsterdam, The Netherlands; Cancer Center Amsterdam, Amsterdam UMC, Amsterdam, The Netherlands; Department of Hematology, Leiden UMC, Leiden, The Netherlands; Department of Infectious Diseases, Amsterdam UMC location University of Amsterdam, Amsterdam, The Netherlands; Department of Hematopoiesis, Sanquin Research, Amsterdam, The Netherlands; Department of Internal Medicine-Hematology, St. Antonius Hospital, Nieuwegein, The Netherlands; Department of Clinical Chemistry, Amsterdam UMC, Amsterdam, The Netherlands; Laboratory Medical Immunology, Department of Immunology, Erasmus MC, University Medical Center, Rotterdam, The Netherlands

**Author notes:** Correspondence:* Prof. dr. M.D. Hazenberg, Department of Hematology, Amsterdam University Medical Center, location AMC, Meibergdreef 9, 1105 AZ Amsterdam, The Netherlands. These authors contributed equally (Sabine Haggenburg, Quincy Hofsink, Abraham Goorhuis, Caroline E. Rutten, Mette D. Hazenberg, Inger S. Nijhof).

## Abstract

**Importance:** In patients with hematologic malignancies, the immunogenicity of the standard 2-dose mRNA-1273 coronavirus disease 19 (COVID-19) vaccination schedule is often insufficient due to underlying disease and current or recent therapy.

**Objective:** To determine whether a 3^rd^ mRNA-1273 vaccination raises antibody concentrations in immunocompromised hematology patients to levels obtained in healthy individuals after the standard 2-dose mRNA-1273 vaccination schedule.

**Design:** Prospective observational cohort study.

**Setting:** Four academic hospitals in the Netherlands.

**Participants:** 584 evaluable immunocompromised hematology patients, all grouped in predefined cohorts spanning the spectrum of hematologic malignancies.

**Exposure:** One additional vaccination with mRNA-1273 5 months after completion of the standard 2-dose mRNA-1273 vaccination schedule.

**Main Outcomes and Measures:** Serum IgG antibodies to spike subunit 1 (S1) antigens prior to and 4 weeks after each vaccination, and pseudovirus neutralization of wildtype, delta and omicron variants in a subgroup of patients.

**Results:** In immunocompromised hematology patients, a 3^rd^ mRNA-1273 vaccination led to median S1 IgG concentrations comparable to concentrations obtained by healthy individuals after the 2-dose mRNA-1273 schedule. The rise in S1 IgG concentration after the 3^rd^ vaccination was most pronounced in patients with a recovering immune system, but potent responses were also observed in patients with persistent immunodeficiencies. Specifically, patients with myeloid malignancies or multiple myeloma, and recipients of autologous or allogeneic hematopoietic cell transplantation (HCT) reached median S1 IgG concentrations similar to those obtained by healthy individuals after a 2-dose schedule. Patients on or shortly after rituximab therapy, CD19-directed chimeric antigen receptor T cell therapy recipients, and chronic lymphocytic leukemia patients on ibrutinib were less or unresponsive to the 3^rd^ vaccination. In the 27 patients who received cell therapy between the 2^nd^ and 3^rd^ vaccination, S1 antibodies were preserved, but a 3^rd^ mRNA-1273 vaccination did not significantly enhance S1 IgG concentrations except for multiple myeloma patients receiving autologous HCT. A 3^rd^ vaccination significantly improved neutralization capacity per antibody.

**Conclusions and Relevance:** The primary schedule for immunocompromised patients with hematologic malignancies should be supplemented with a delayed 3^rd^ vaccination. B cell lymphoma patients and allogeneic HCT recipients need to be revaccinated after treatment or transplantation.

**Trial Registration:** EudraCT 2021-001072-41

**Key points:** *Question:* Can a 3^rd^ mRNA-1273 vaccination improve COVID-19 antibody concentrations in immunocompromised hematology patients to levels similar to healthy adults after the standard 2-dose mRNA-1273 schedule?

*Findings:* In this prospective observational cohort study that included 584 immunocompromised hematology patients, a 3^rd^ mRNA-1273 vaccination significantly improved SARS-CoV-2 antibody concentrations to levels not significantly different from those obtained by healthy individuals after the standard 2-dose mRNA-1273 vaccination schedule. Pseudovirus neutralization capacity per antibody of wild type virus and variants of concern also significantly improved.

*Meaning:* The primary COVID-19 vaccination schedule for immunocompromised patients with hematologic malignancies should be supplemented with a delayed 3^rd^ vaccination.

## Introduction

Patients with hematologic diseases are at high risk for severe coronavirus disease 2019 (COVID-19), COVID-19-related death, and persistent viral shedding, also in the omicron era.^1–5^ While vaccination has demonstrated efficacy against COVID-19-related hospital admission and death in healthy individuals,^6–8^ several studies among COVID-19 vaccinated hematology patients demonstrated reduced severe acute respiratory syndrome coronavirus 2 (SARS-CoV-2) seroconversion rates^9–16^ and lowered COVID-19 vaccine effectiveness.^17,18^ Reduced vaccine immunogenicity in hematology patients is related to the disease itself and the therapy thereof.^19,20^

We prospectively measured antibody responses to mRNA-1273 (Moderna/Spikevax) vaccination in those hematology patients that are generally considered too immunocompromised to mount an effective immune response and in whom vaccinations are often postponed until (later) after treatment.^12,21,22^ In collaboration with a number of other prospective cohort studies on the immunogenicity of COVID-19 vaccination among immunocompromised and healthy individuals conducted in the Netherlands we quantified antibody concentrations against the WHO standard and set a spike-1 (S1) IgG concentration of 300 BAU/ml as the lower threshold of an adequate COVID-19 vaccine response.^23–28^ This is in line with an independent British cohort where a lower threshold of 264 BAU/ml was estimated to correspond to a vaccine efficacy of 80% against symptomatic SARS-CoV-2 infection.^29^ More than half of the patients in our study (55%) obtained an S1 IgG antibody concentration ≥300 BAU/ml after the standard 2-dose mRNA-1273 vaccination schedule, despite their immunodeficiencies. Many patients who remained below this threshold after completion of the standard 2-dose mRNA-1273 schedule nevertheless showed an increase in SARS-CoV-2 antibody concentration with each vaccination.^12^ This raised the question whether a 3^rd^ vaccination could further improve SARS-CoV-2 immunity. After early reports on waning immunity in healthy individuals,^28,30^ and with the surge of the SARS-CoV-2 delta variant of concern, immunocompromised individuals were offered a 3^rd^ mRNA-1273 vaccination in the fall of 2021. The goal of this study was to determine whether this 3^rd^ vaccination could enhance antibody concentrations in immunocompromised hematology patients to levels obtained in healthy individuals after the standard 2-dose mRNA-1273 schedule.

## Methods

### Study participants

In this prospective, observational multicenter cohort study we analyzed antibody responses to a 3-dose mRNA-1273 vaccination schedule in 723 adult hematology patients, classified into pre-defined cohorts based on their diagnosis and treatment status at the start of the study (Table 1). Patient characteristics, in- and exclusion criteria are described in detail elsewhere.^12^ In the present analysis sickle cell patients were excluded as they did not receive a 3^rd^ vaccination at the same time as the patients with hematologic malignancies, according to the Dutch COVID-19 vaccination protocol. All other participants were offered a 3^rd^ dose 5 months after completion of the standard 2-dose mRNA-1273 vaccination schedule (Figure 1A). Study protocols were approved by the Institutional Review Board of the Amsterdam UMC location Free University and participating centers. All patients provided written informed consent prior to study onset.

**Table 1.** Patient characteristics stratified by cohort. Indicated treatment as per first mRNA-1273 vaccination. ^A^S1 IgG>10 BAU/ml; ^B^2^nd^ vaccination vs healthy control; ^C^3^rd^ vaccination vs healthy control after 2^nd^ vaccination. Mo: months. BEAM: BCNU, etoposide, cytarabine, melphalan; IMiDs: immunomodulatory imide drugs; HCT: hematopoietic progenitor cell transplantation; CLL: chronic lymphocytic leukemia; CML: chronic myeloid leukemia; AML: acute myeloid leukemia; MDS: myelodysplastic syndrome; GvHD: graft versus host disease; CAR: chimeric antigen receptor.

|  | N | Age | Sex | Seroconversion<sup>A</sup> |  | S1 IgG concentration (BAU/ml) |  |  |  |
| --- | --- | --- | --- | --- | --- | --- | --- | --- | --- |
|  |  | Mean<br>(SD) | Women<br>(%) | After 2<sup>nd</sup><br>n (%) | After 3<sup>rd</sup><br>n (%) | After 2<sup>nd</sup><br>(median) | p-<br>value<sup>B</sup> | After 3<sup>rd</sup><br>(median) | p-<br>value<sup>C</sup> |
| **All patients** | 584 | 60 (11.2) | 216<br>(37.0) | 399<br>(68.9) | 443<br>(78.8) | 303.1 | 0.00 | 2171.3 | *0.94 |
| **Lymphoma** |  |  |  |  |  |  |  |  |  |
| During rituximab ± chemotherapy | 40 | 59 (13.0) | 16 (40.0) | 6 (15.0) | 9 (22.5) | 0.7 | 0.00 | 0.7 | 0.00 |
| <12mo after rituximab ±<br>chemotherapy | 36 | 62 (11.0) | 16 (44.4) | 14 (40.0) | 24 (70.6) | 4.2 | 0.00 | 292.5 | 0.01 |
| <12mo after autologous HCT (BEAM) | 25 | 59 (12.0) | 9 (36.0) | 13 (52.0) | 14 (58.3) | 15.4 | 0.00 | 731.0 | *0.26 |
| **Multiple myeloma** |  |  |  |  |  |  |  |  |  |
| 1<sup>st</sup> line therapy | 23 | 62 (8.0) | 9 (39.1) | 17 (77.3) | 20 (95.2) | 435.1 | 0.06 | 411.2 | *0.55 |
| Daratumumab | 44 | 63 (8.0) | 17 (38.6) | 42 (95.5) | 43 (97.7) | 539.3 | 0.00 | 1729.0 | *0.96 |
| IMiDs | 46 | 60 (8.0) | 17 (37.0) | 41 (89.1) | 40 (88.9) | 1064.2 | *0.17 | 3071.8 | *0.17 |
| <9mo after autologous HCT (HDM) | 44 | 61 (7.0) | 12 (27.3) | 42 (95.5) | 43 (97.7) | 2457.1 | *0.44 | 10589.4 | ^0.00 |
| **CLL** |  |  |  |  |  |  |  |  |  |
| Watch & wait | 43 | 64 (8.0) | 19 (44.2) | 34 (81.0) | 34 (85.0) | 535.4 | 0.01 | 3465.0 | *0.27 |
| Ibrutinib | 33 | 64 (7.0) | 12 (36.4) | 12 (36.4) | 18 (54.5) | 1.3 | 0.00 | 3.9 | 0.00 |
| **CML** |  |  |  |  |  |  |  |  |  |
| Tyrosine kinase inhibitor | 42 | 54 (14.0) | 17 (40.5) | 42<br>(100.0) | 39<br>(100.0) | 2658.7 | *0.31 | 7572.1 | ^0.00 |
| **AML and high-risk MDS** |  |  |  |  |  |  |  |  |  |
| Hypomethylating therapy | 16 | 66 (14.0) | 3 (18.8) | 12 (75.0) | 14 (93.3) | 115.7 | 0.00 | 836.3 | *0.16 |
| High-dose chemotherapy | 18 | 52 (15.0) | 8 (44.4) | 16 (94.1) | 18<br>(100.0) | 3795.9 | *0.11 | 11379.1 | ^0.00 |
| **Myeloproliferative disease** |  |  |  |  |  |  |  |  |  |
| Ruxolitinib | 31 | 58 (11.0) | 13 (41.9) | 30 (96.8) | 28 (96.6) | 296.2 | 0.00 | 1795.9 | *0.73 |
| **Allogeneic HCT** |  |  |  |  |  |  |  |  |  |
| < 6 mo after HCT | 49 | 55 (13.0) | 19 (38.8) | 28 (57.1) | 39 (84.8) | 20.9 | 0.00 | 2993.9 | *0.50 |
| Chronic GvHD | 51 | 57 (9.0) | 17 (33.3) | 41 (80.4) | 46 (92.0) | 1064.6 | 0.04 | 4830.4 | *0.07 |
| **CAR T cell therapy** |  |  |  |  |  |  |  |  |  |
| CD19-directed | 43 | 60 (12.0) | 12 (27.9) | 9 (21.4) | 14 (35.0) | 0.3 | 0.00 | 0.5 | 0.00 |
| **Healthy control** |  |  |  |  |  |  |  |  |  |
| PIENTER cohort | 44 | 57 (10) | 30 (68) | 42 (95.5) | n.a. | 1965.4 | - | n.a. | - |

**Figure 1.**
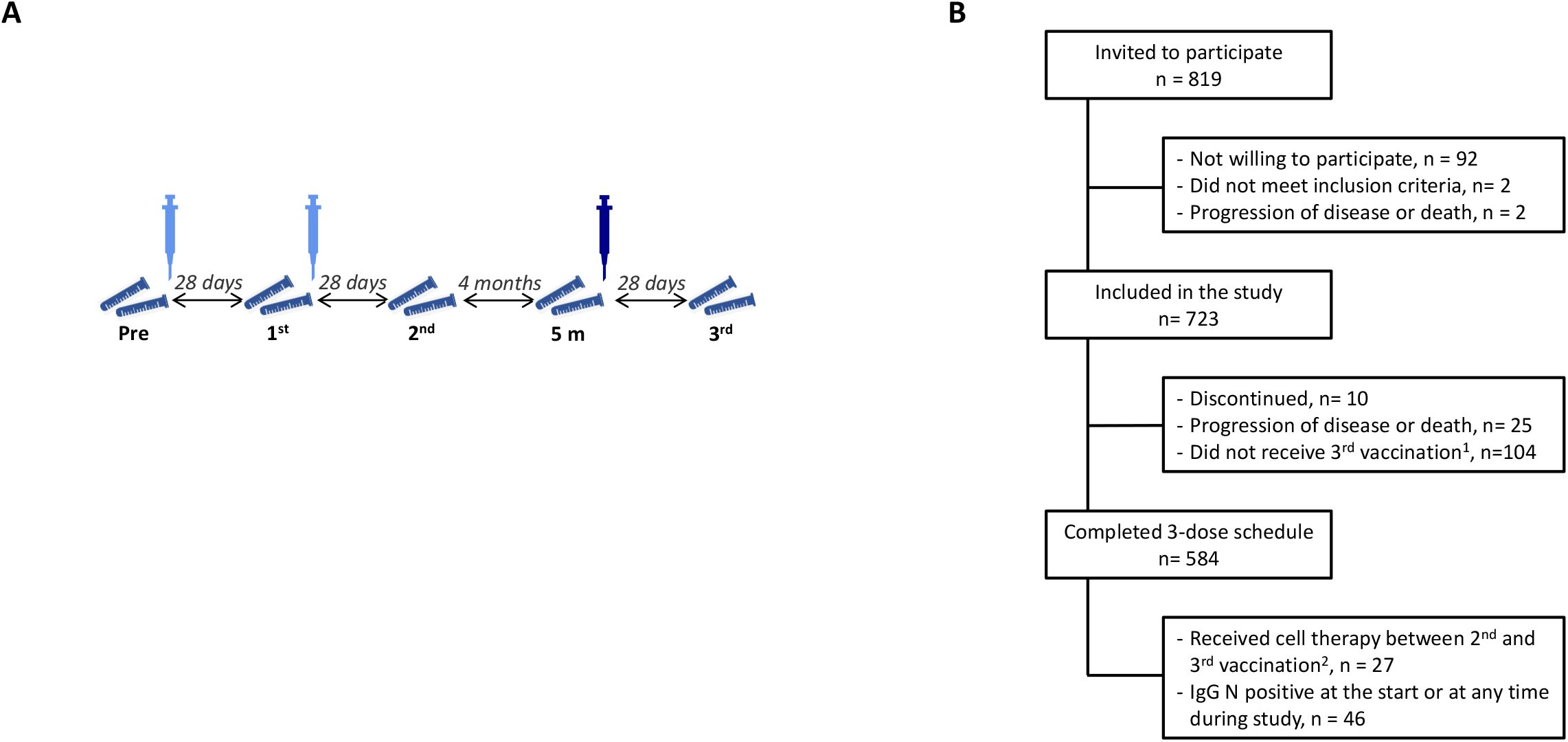
Study protocol and patients. **A**. Synopsis of study protocol. Syringe indicates mRNA-1273 vaccination, tubes indicate collection of blood for analysis of the effect of the 1^st^, 2^nd^ and 3^rd^ vaccination. **B**. Patient inclusion. Details on ^1^patients that did not receive a 3^rd^ vaccination during the time under study are depicted in eTable 2, and on ^2^patients that received cell therapy between the 2^nd^ and 3^rd^ vaccination in eTable 4.

### Clinical parameters

B, T and NK cell numbers, demographic parameters and medical history including comorbidities and concomitant medication (via standardized case report forms) were collected prior to and 28 days after each vaccination.

### Antibody concentrations and neutralization

Humoral responses against S1, receptor binding domain (RBD) and nucleocapsid (N) antigen domains of SARS-CoV-2 were quantified 28 days after each vaccination as described.^12,31^ SARS-CoV-2 binding antibody concentration was calibrated against the National Institute for Biological Standards Control (NIBSC) first serum standard for COVID-19 (20/136), as recommended by the World Health Organization (WHO).^23,29,31^ Seroconversion was defined as obtaining an S1 IgG concentration >10 binding antibody units (BAU)/ml, and an adequate vaccine response as S1 IgG ≥300 BAU/ml, the IgG concentration that met a SARS-CoV-2 wildtype (Wuhan) virus PRNT50 (plaque reduction neutralization titer) of 40 or higher in 2 independent prospective Dutch mRNA-1273 vaccination cohorts.^25,26,32,33^ Reference antibody levels were extracted from randomly selected age-matched Dutch citizens who had received a 2^nd^ dose of mRNA-1273 14-61 (median 49) days prior to blood sampling (PIENTER cohort^12,27^). Antibody neutralization activity was tested using lentiviral-based pseudoviruses expressing the SARS-CoV-2 wild type (D614G), delta (B.1.617.2) and omicron (BA.1) variants.^34^ We determined the 50% inhibitory dose (ID50) in all low-responder patients (S1 IgG 50-300 BAU/ml) and in a random selection of adequate responder patients (S1 IgG ≥300 BAU/ml) after the 2-dose mRNA-1273 schedule (eTable 1).^12,28^

### Statistical analysis

Differences between groups and timepoints were analyzed with Mann-Whitney U and paired sample t-tests after ^10^log-transformation, respectively. Pearson correlation was calculated between serum S1 IgG and pseudovirus neutralization after ^10^log-transformation of both. S1 IgG antibody concentrations <300 or ≥300 BAU/ml after 2^nd^ and after the 3^rd^ vaccination were compared with the McNemar test. Analyses with two-sided p-values of <0.05 were considered statistically significant. Statistical analyses were performed using the IBM SPSS Statistics for Windows, Version 26.0 (IBM Corp., Armonk, NY) and R for Windows, Version 4.0.3 (The R Foundation for Statistical Computing, Vienna, Austria).

## Results

### SARS-CoV-2 antibody concentration before and after the 3^rd^ mRNA-1273 vaccination

Of 723 study participants, 584 patients completed the 3-dose mRNA-1273 schedule (Figure 1b; Table 1). Most of the 104 patients who did not receive a 3^rd^ vaccination deferred the 3^rd^ dose because they felt sufficiently protected after 2 mRNA-1273 vaccinations (median S1 IgG 2323 BAU/ml; eTable 2). Twenty-four patients (4.1%) had been infected with SARS-CoV-2 prior to the first vaccination as identified by the presence of nucleocapsid antibodies (N IgG > 14.3 BAU/ml),^35^ and 22 patients (3.8%) became infected during follow up (eTable 3, eFigure 1). Twenty-seven patients (4.6%) received cell therapy at any time during the 5 months between the 2^nd^ and the 3^rd^ vaccination (eTable 4). SARS-CoV-2 infected and intercurrent cell therapy patients were analyzed separately.

Of evaluable participants, 50.2% obtained S1 IgG antibody concentrations ≥300 BAU/ml after completion of the standard 2-dose vaccination schedule, while 32.7% did not seroconvert (S1 IgG <10 BAU/ml; Figure 2a). Median S1 IgG concentration of patients (303.1 BAU/ml) remained significantly lower than median S1 IgG concentration of age-matched healthy controls (1965.4 BAU/ml; p<0.001) after the 2-dose mRNA-1273 schedule. In the 5 months interval between the 2^nd^ and 3^rd^ vaccination, S1 IgG concentrations declined significantly to a median of 92.5 BAU/ml, with only 30.2% of patients maintaining S1 IgG ≥300 BAU/ml (Figure 2a). A 3^rd^ mRNA-1273 vaccination led to a significant increase in S1 IgG concentration. The majority of patients (78.8%) seroconverted and 65.9% obtained S1 IgG ≥300 BAU/ml. Median S1 IgG concentration after the 3^rd^ mRNA-1273 vaccination in patients (2171.3 BAU/ml) was no longer significantly lower than the concentration obtained by healthy individuals after the standard 2-dose mRNA-1273 schedule (p=0.46; Figure 2a). The majority (72.6%) of low-responder patients (S1 IgG 10-300 BAU/ml) after the 2^nd^ vaccination obtained an S1 IgG concentration ≥300 BAU/ml after the 3^rd^ vaccination. Serum RBD IgG concentrations demonstrated similar dynamics (eFigure 2).

**Figure 2.**
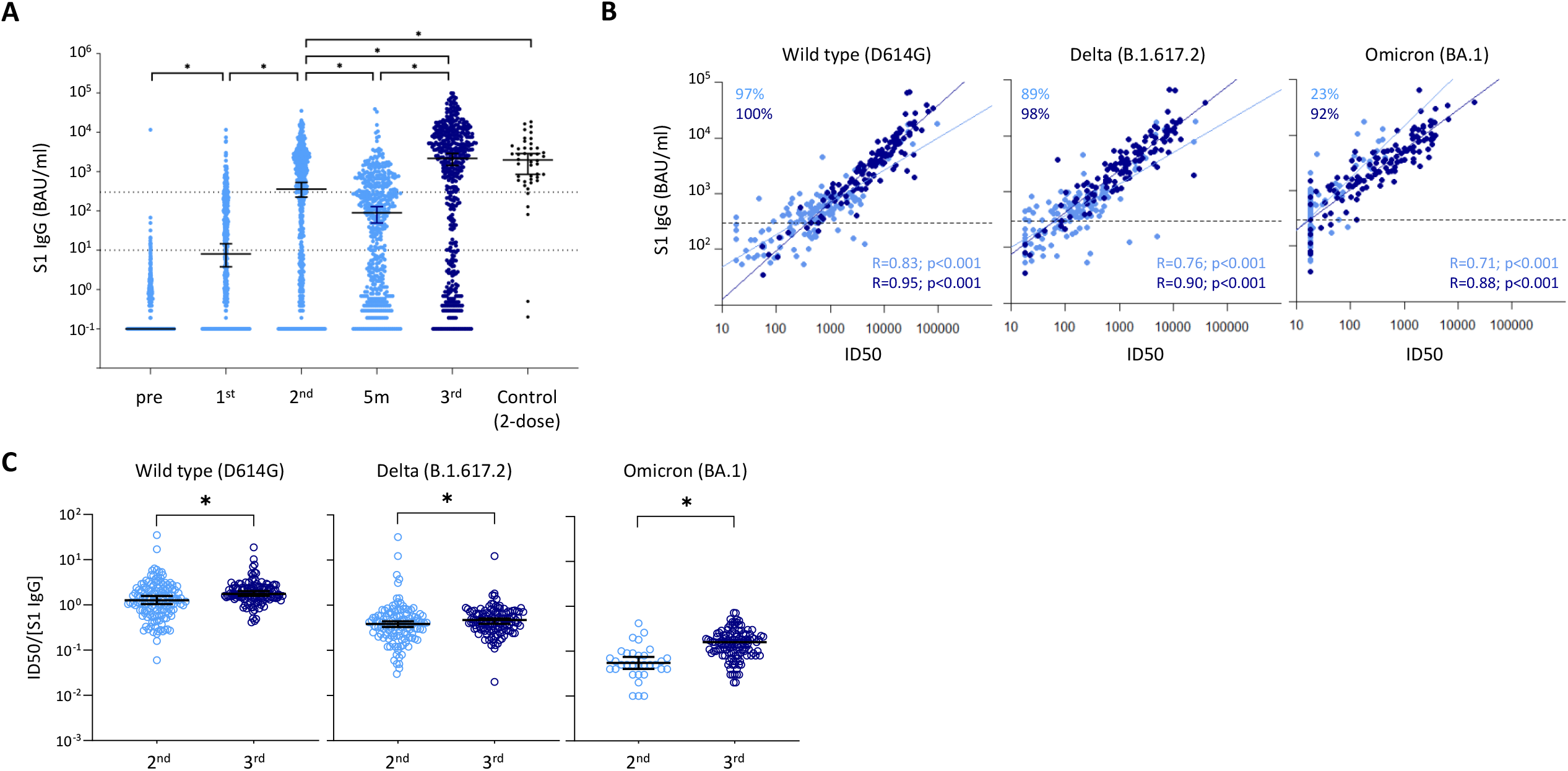
S1 binding antibody concentration and neutralization. **A**. IgG S1 concentration at each timepoint for previously uninfected patients. IgG S1 concentration of age-matched controls measured after the standard 2-dose mRNA-1273 schedule is indicated in black. Dotted lines: seroconversion (S1 IgG >10 BAU/mL) and S1 IgG concentration ≥300 BAU/ml. **B**. Correlation between IgG S1 concentration and pseudovirus neutralization for SARS-CoV-2 wild type virus and variants of concern for all patients with S1 IgG 10-300 BAU/ml and a subset of patients with S1 IgG ≥300 BAU/ml after the 2^nd^ vaccination (eTable 1; reference 12). Indicated in the top-left corner are percentages of patients with ID50 >20 (the lower limit of quantification). **C**. Pseudovirus neutralization ID50 / S1 IgG concentration ratio of patients with ID50 >20 for wildtype SARS-CoV-2 and variants of concern. Light blue: standard 2-dose mRNA schedule; dark blue: additional 3^rd^ mRNA-1273 vaccination. Asterix: p<0.05.

### Improvement of neutralization capacity

Serum S1 IgG concentration correlated significantly with pseudovirus neutralization of SARS-CoV-2 wild type (r=0.83; p<0.001) and variants of concern (delta: r=0.76; p<0.001; omicron BA.1: r=0.71; p<0.001) after the 2^nd^ vaccination, and even stronger after the 3^rd^ vaccination (wild type: r=0.95; p<0.001; delta: r=.0.90; p<0.001; omicron BA.1: r=0.88; p<0.001) (Figure 2b). The ratio between pseudovirus neutralization and S1 IgG concentration can be taken as a measure for antibody neutralization capacity.^36^ Neutralization capacity of wild type SARS-CoV-2, delta and omicron BA.1 variants per S1 IgG antibody improved significantly after the 3^rd^ vaccination (Figure 2c).

### Significant S1 IgG increase in almost all patient cohorts after the 3^rd^ vaccination

A 3^rd^ mRNA-1273 dose led to a significant increase in S1 IgG concentration in all cohorts, with the exception of B cell non-Hodgkin’s lymphoma (B-NHL) patients with ongoing B cell depletion due to rituximab therapy (Figure 3a). The steepest increase in median S1 IgG concentration was observed in patients with a recovering immune system: 36-fold for patients with absent B cells at the time of the primary vaccination schedule (‘<12 months after rituximab’ cohort), 15-fold for B-NHL patients who had received myeloablative chemotherapy and autologous HCT less than 12 months prior to the 1^st^ vaccination, and 49-fold in patients who had received allogeneic HCT less than 6 months before the 1^st^ vaccination. This was confirmed in a paired analysis with S1 IgG concentration <300 or ≥300 BAU/ml as a dichotomous outcome after the 2^nd^ and after the 3^rd^ vaccination (not shown). Significant increases in median S1 IgG concentrations were also observed in patients with ongoing immunosuppression, e.g. patients with acute myeloid leukemia (AML) or high-risk myelodysplastic syndrome (MDS) on hypomethylating therapy, patients with myeloproliferative neoplasms on ruxolitinib and patients with chronic graft versus host disease (GvHD) using oral immunosuppressants (Figure 3a). In fact, in 12 out of the 16 cohorts, the 3-dose schedule induced S1 IgG concentrations to similar or even higher levels as the 2-dose mRNA-1273 schedule did in healthy individuals (Table 1).

**Figure 3.**
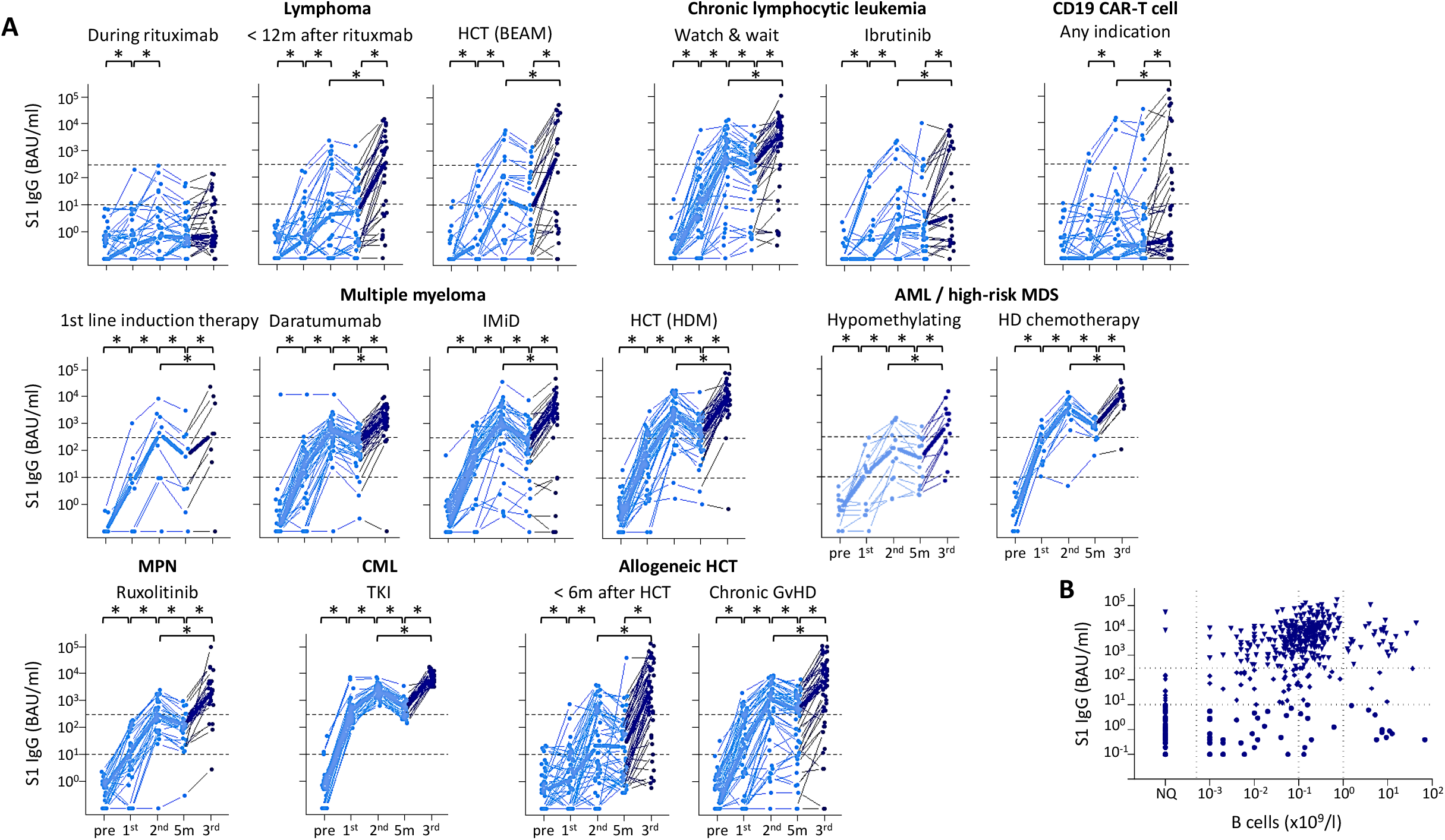
S1 binding antibody concentration for each patient. **A**. IgG S1 concentrations over time of previously uninfected patients grouped per cohort. In light blue 1^st^ and 2^nd^ vaccination, in dark blue 3^rd^ vaccination. Asterix: p<0.05. **B**. S1 IgG concentration 4 weeks after the 3^rd^ vaccination versus B cell number at the time of 3^rd^ vaccination. Dotted lines indicate seroconversion (S1 IgG >10 BAU/mL), S1 IgG concentration ≥300 BAU/ml), upper and lower limits of normal B cell numbers and B cell number detection limit. NQ: not quantifiable.

### Low and non-responder patients

The only cohorts in which median S1 IgG concentrations remained low (<300 BAU/ml) or absent (<10 BAU/ml) were those with patients on, or shortly after, rituximab therapy, with patients who received CD19-directed CAR T cell therapy and CLL patients on ibrutinib (Figure 3a, Table 1). The few CAR T cell recipients who did obtain S1 IgG ≥300 BAU/ml had low to normal B cell numbers at the time of the 3^rd^ vaccination. The majority (84.8%) of patients who did not seroconvert after the 3^rd^ vaccination (S1 IgG <10 BAU/ml) had ongoing B cell depletion, due to CD20 antibody therapy or CD19 directed CAR T cell therapy (Figure 3b). This included 6 of the 7 allogeneic HCT recipients in whom S1 IgG concentrations remained <10 BAU/ml even after the 3^rd^ vaccination. Of non-responder patients, 9.8% were CLL patients with higher-than-normal B cell numbers at time of the 3^rd^ vaccination. However, higher-than-normal B cell numbers did not preclude S1 IgG concentrations ≥300 BAU/ml after the 3^rd^ vaccination (Figure 3b). There were no other common denominators but absence of B cells to predict low (S1 IgG 10-300 BAU/ml) or non-responder status after the 3^rd^ vaccination.

### Impact of HCT or CAR T cell therapy between the 2^nd^ and 3^rd^ vaccination

Twenty-seven patients received cell therapy (CAR T cell therapy or HCT) after completion of the primary 2-dose schedule, and before receiving the 3^rd^ mRNA-1273 vaccination (eTable 4, Figure 4). All patients demonstrated a similar significant decline in S1 IgG concentration in the 5 months between the 2^nd^ and the 3^rd^ vaccination as observed in the other cohorts (Figure 3a), but in none of the patients cell therapy lead to S1 IgG concentrations <10 BAU/ml (Figure 4). Multiple myeloma patients who received high-dose melphalan (HDM) followed by autologous HCT after the 2^nd^ vaccination demonstrated a significant increase in S1 IgG concentration after the 3^rd^ vaccination (Figure 4). A similar S1 IgG dynamic was observed in the one patient who received busulfan/cyclophosphamide followed by autologous HCT after the 2^nd^ vaccination as consolidation therapy for AML. In the few B-NHL patients who received autologous HCT or CAR T cell therapy after the 2^nd^ vaccination a 3rd mRNA-1273 dose did not increase S1 IgG concentrations. Similarly, in patients who received an allogeneic HCT after the 2^nd^ vaccination S1 IgG concentrations did not increase after the 3^rd^ dose (Figure 4). With the exception of 1 patient, all new allogeneic HCT recipients had B cell numbers <0.1×10^9^/ml and used immunosuppressants (5 patients ≥2) at the time of the 3^rd^ dose (eTable 4).

**Figure 4.**
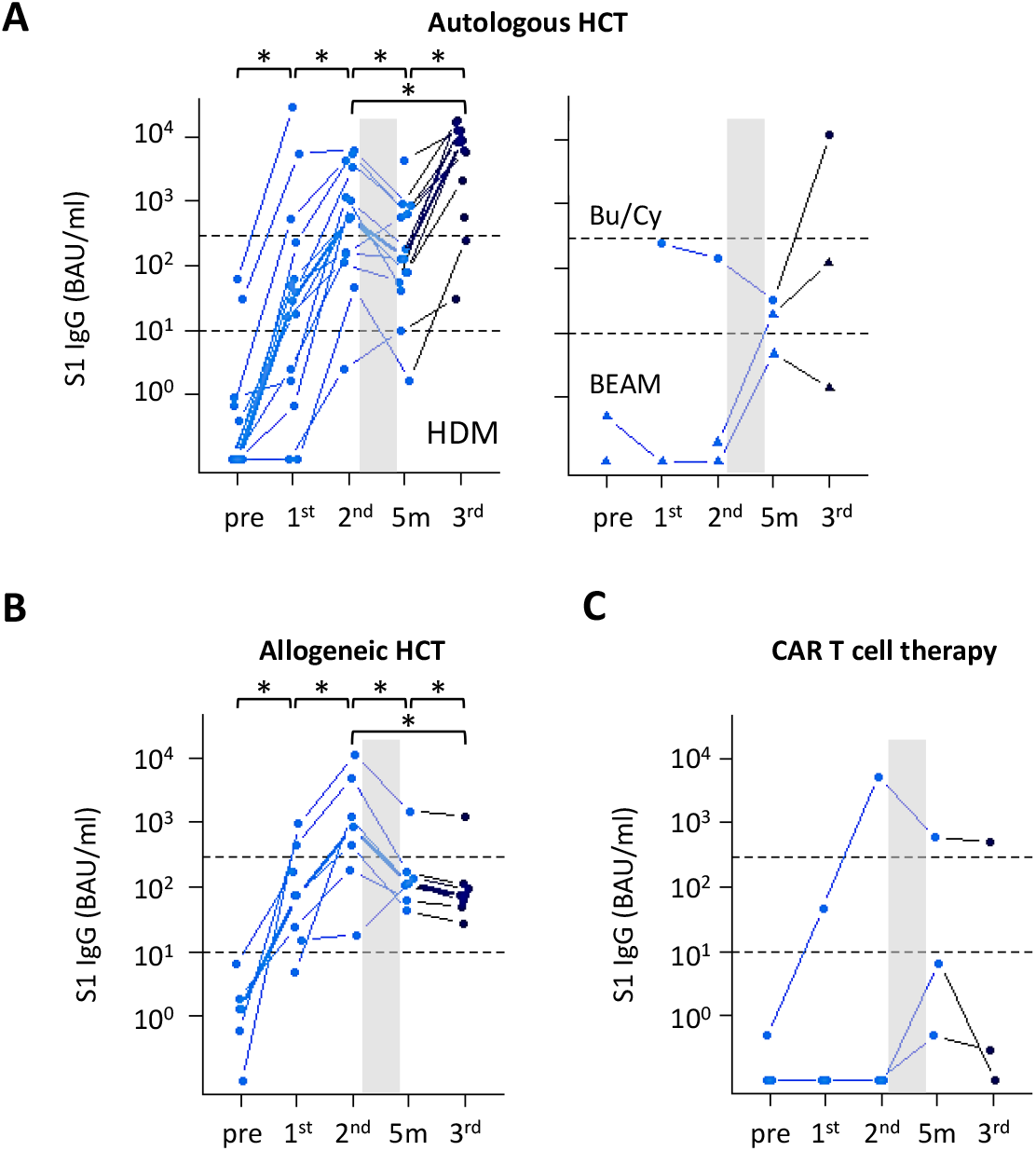
IgG S1 concentration in patients who received cell therapy after the 2^nd^ vaccination. Patients who received cell therapy as indicated (grey bar) at any time during the 5 months between the 2^nd^ and the 3^rd^ mRNA-1273 vaccination. See for patient details eTable 4. **A**. Autologous HCT, left panel: multiple myeloma patients who received the 1^st^ and 2^nd^ vaccination during 1^st^ line remission-induction therapy and the 3^rd^ vaccination at a median of 102 days after high-dose melphalan (HDM) and autologous HCT. Autologous HCT, right panel: AML patient who received busulphan/cyclophosphamide (Bu/Cy) followed by autologous HCT (circles), and B-NHL patients who received BEAM and autologous HCT (triangles). **B**. Patients who received an allogeneic HCT between the 2^nd^ and 3^rd^ vaccination. **C**. Patients who received CAR T cells between the 2^nd^ and 3^rd^ vaccination. Dotted lines: seroconversion (S1 IgG > 10 BAU/mL) and S1 IgG concentration ≥300 BAU/ml. Bold lines in left panels indicate median values; asterix: p<0.05; number of patients in right panel in A and in C was too low for statistical analyses.

### SARS-CoV-2 infection

Nine patients became infected with SARS-CoV-2 in the 5 months between the 2^nd^ and 3^rd^ vaccination, a time when first the alpha and later the delta SARS-CoV-2 variant was most prevalent in the Netherlands. Infection was confirmed by a positive PCR test and by the presence of N IgG serum antibodies (>14.3 BAU/ml).^35^ Five patients were asymptomatic, 3 of them had developed S1 IgG ≥300 BAU/ml after the 2^nd^ vaccination, 1 was a low responder (S1 IgG 10-300 BAU/ml) and 1 was a non-responder (S1 IgG <10 BAU/ml) after the 2^nd^ vaccination. The other four had symptomatic infection, of whom 3 were non-responders (S1 IgG <10 BAU/ml after the 2^nd^ vaccination); 1 of these non-responders was admitted to the intensive care unit. Of the 7 patients who became SARS-CoV-2 infected after the 3^rd^ vaccination, only 1 reported mild symptoms. None of the study participants died of COVID-19.

## Discussion

In this study we demonstrate that with a 3^rd^ mRNA-1273 vaccination, the majority of immunocompromised hematology patients obtained SARS-CoV-2 antibody concentrations similar to healthy individuals after the standard 2-dose mRNA-1272 schedule. Whether a 3-dose mRNA-1273 schedule offers similar protection against severe COVID-19 in immunocompromised hematology patients as the standard 2-dose regimen in healthy individuals remains to be determined. We did observe a significant correlation between S1 IgG concentration and neutralization of SARS-CoV-2 wild type, delta and omicron BA.1 variants of concern. Neutralization capacity per antibody unit improved significantly after the 3^rd^ vaccination, suggesting antibody maturation over time.^36^ This is similar to what has been observed in healthy individuals where it was related to improved antibody avidity.^36^ Antibody maturation occurred across study cohorts, despite for example the depletion of plasma cells in daratumumab treated multiple myeloma patients or the broad immunodeficiency observed in jak-2 inhibitor treated patients or allogeneic HCT recipients.

Only in CLL patients on ibrutinib^37^ and in patients with ongoing B cell depletion, most often due to continued CD20 antibody therapy or CD19-directed CAR T cell therapy, antibody responses remained low. This does not preclude the generation of potent T cell responses, as was demonstrated recently in patients with rheumatologic or hematologic conditions.^38–40^ As T cell responses have been demonstrated to protect B cell depleted hematology patients against severe COVID-19,^41^ boosting T cell responses with a 3^rd^ vaccination may, by inference, further harness patients against severe COVID-19. This, however, remains to be confirmed.

The additional value of a 3^rd^ vaccination was most pronounced in patients in whom the immune system had recovered in the time after receiving the primary 2-dose vaccination schedule. In B-NHL patients, SARS-CoV-2 antibodies appeared in parallel with recovering B cell numbers. Importantly, and as reported previously,^12^ B cells did not need to reach normal values to produce adequate S1 IgG concentrations. In CD19-directed CAR T cell patients, adequate antibody concentrations were obtained in those patients in whom B cells had re-appeared. In allogeneic HCT recipients, reconstitution of the lymphocyte pool in the months after transplantation, in combination with tapering of immunosuppressants, led to pronounced increases in S1 IgG concentrations after the 3^rd^ vaccination.^42^ Importantly, also in patients with ongoing immunodeficiency at the time of the 3^rd^ mRNA-1273 dose, S1 IgG concentrations significantly increased. Multiple myeloma and untreated CLL are associated with immunodeficiencies that hamper vaccination-induced antibody responses.^20^ Nevertheless, a 3^rd^ mRNA-1273 vaccination in these patients led to a significant increase in S1 IgG concentrations, to levels that were no longer significantly different from those obtained in healthy individuals after the standard 2-dose mRNA-1273 schedule. Similar dynamics were observed in patients with myeloproliferative neoplasms on the JAK-2 inhibitor ruxolitinib, patients with AML on hypomethylating therapy and patients with chronic GvHD. While in none of these patient groups the underlying immunodeficiency had been resolved, a 3^rd^ dose of mRNA-1273 vaccination brought antibody concentrations to adequate levels.

It is often thought that high-dose chemotherapy and autologous HCT undo previously generated immunity. It is for this reason that almost all international vaccination protocols offer an intensive revaccination program from at least 6 months after transplantation. We demonstrated previously that COVID-19 vaccine immunogenicity depends strongly on the underlying hematologic disease.^12^ Vaccination did not yield adequate SARS-CoV-2 antibody concentrations in B-NHL patients in the first 8 months after autologous HCT, or in the first 4-6 months after allogeneic HCT. By contrast, in multiple myeloma patients HDM and autologous HCT did not hamper strong antibody responses, not even when patients were vaccinated as early as 2-4 weeks after autologous HCT.^12^ In line with these observations, we demonstrate that COVID-19 vaccinated B-NHL patients who proceed to autologous HCT should be offered a SARS-CoV-2 revaccination schedule after transplant, while this is not necessary for COVID-19 vaccinated multiple myeloma patients. Patients who received allogeneic HCT after the primary COVID-19 vaccination schedule should be revaccinated after transplantation. Our data suggest however that not all acquired immunity is lost, and the number and timing of revaccinations after transplantation remain to be determined.

Together these data indicate that with a 3^rd^ mRNA-1273 vaccination added to the standard 2-dose schedule, the majority of hematology patients with recovering immunity but also patients with ongoing immunodeficiencies obtain SARS-CoV-2 antibody concentrations that are not significantly different from concentrations obtained in healthy individuals after the standard 2-dose vaccination schedule. To compensate for reduced in vitro neutralization of variants of concern such as the omicron BA-2 variant, booster vaccinations are indicated on top of the 3-dose mRNA-1273 schedule, similar as in healthy individuals. It is possible that the most immunocompromised patients may need 2 or more booster vaccinations. COVID-19 vaccination in immunocompromised patients should be based on a primary 3-dose mRNA-1273 schedule instead of the 2-dose schedule that is standard for healthy individuals.

## Data Availability

All data produced in the present study are available upon reasonable request to the authors.

## Acknowledgements

The authors would like to extend their deepest gratitude to all patients, colleagues, students and volunteers from participating institutes, the Netherlands Cancer Registry and the National Institute for Public Health and the Environment who made this study possible. This study was financially supported by the Dutch Research Council (NWO ZonMW, grant 10430072010009) and Amsterdam UMC.

## Notes

**Potential conflict of interest:** J.C. is consultant for Novartis, received research funding from Novartis, Merus, Takeda, Genentech, BD Biosciences and royalties from Navigate and BD Biosciences. A.P.K. is consultant for Abbvie, Janssen, Astra Zeneca, BMS, Roche, LAVA, received speakers fees from Abbvie and research funding from Janssen, Abbvie, Astra Zeneca, BMS, Celgene and Roche. S.Z. received research funding from Takeda and Janssen and participated advisory boards of Takeda, Janssen, BMS/Celgene, Sanofi and Oncopeptides. T.M. is consultant for Janssen and Kite, received research funding and honoraria from BMS/Celgene, Genentech and Kite. P.M. received research funding from Astra Zenica and is consultant for GSK and BMS. I.S.N. is consultant for Janssen, BMS/Celgene, Amgen and Sanofi. None of these are conflicts of interest in the present study.

### Competing Interest Statement

Potential conflict of interest: J.C. is consultant for Novartis, received research funding from Novartis, Merus, Takeda, Genentech, BD Biosciences and royalties from Navigate and BD Biosciences. A.P.K. is consultant for Abbvie, Janssen, Astra Zeneca, BMS, Roche, LAVA, received speakers fees from Abbvie and research funding from Janssen, Abbvie, Astra Zeneca, BMS, Celgene and Roche. S.Z. received research funding from Takeda and Janssen and participated advisory boards of Takeda, Janssen, BMS/Celgene, Sanofi and Oncopeptides. T.M. is consultant for Janssen and Kite, received research funding and honoraria from BMS/Celgene, Genentech and Kite. P.M. received research funding from Astra Zenica and is consultant for GSK and BMS. I.S.N. is consultant for Janssen, BMS/Celgene, Amgen and Sanofi. None of these are conflicts of interest in the present study.

### Clinical Trial

EudraCT 2021-001072-41

### Author Declarations

Ethics committee of the Amsterdam UMC, location Vrije University.

