## supplemental tables for "Three-dose mRNA-1273 vaccination schedule: sufficient antibody response in majority of immunocompromised hematology patients"

### eTable 1

| | After 2-dose mRNA-1273 | | | | After 3-dose mRNA-1273 | | | |
|---|---|---|---|---|---|---|---|---|
| | n | Age mean (SD) | Sex women (%) | B cell ($x10^9$/l) [1] median (range) | n | Age mean (SD) | Sex women (%) | B cell ($x10^9$/l) [2] median (range) |
| **All patients** | 161 | 60.2 (10) | 64 (39.8) | 0.10 (0.0-86.0) | 129 | 60.1 (10.3) | 53 (41.1) | 0.11 (0.00-14.95) |
| **Lymphoma** | | | | | | | | |
| During rituximab +/- chemotherapy | 2 | 70 (6) | 0 (0) | 0.00 (0.00-0.00) | 1 | 74 (0) | 0 (0.0) | 0.00 (0.00-0.00) |
| < 12 months after rituximab +/- chemotherapy | 6 | 60 (15) | 2 (33.3) | 0.01 (0.00-0.04) | 4 | 61 (17) | 2 (50.0) | 0.15 (0.00-0.45) |
| < 12 months after autologous HCT (BEAM) | 7 | 53 (17) | 4 (57.1) | 0.12 (0.00-0.19) | 5 | 50 (19) | 4 (80.0) | 0.22 (0.17-0.23) |
| **Multiple myeloma** | | | | | | | | |
| 1st line therapy | 7 | 58 (9) | 3 (42.9) | 0.03 (0.01-0.04) | 2 | 68 (4) | 1 (50.0) | 0.04 (0.01-0.08) |
| Daratumumab-containing therapy | 28 | 63 (7) | 12 (40.0) | 0.05 (0.00-0.62) | 30 | 63 (7) | 12 (40.0) | 0.04 (0.00-0.33) |
| IMiDs | 15 | 60 (7) | 5 (33.3) | 0.10 (0.00-0.27) | 14 | 59 (6) | 5 (35.7) | 0.09 (0.00-0.25) |
| < 9 months after autologous HCT (HDM) | 13 | 62 (7) | 4 (30.8) | 0.12 (0.01-86) | 10 | 61 (7) | 3 (30.0) | 0.11 (0.03-0.37) |
| **Chronic lymphocytic leukemia** | | | | | | | | |
| Wait & see | 14 | 63 (9) | 6 (42.9) | 8.31 (1.10-75.01) | 12 | 62 (9) | 5 (41.6) | 7.50 (1.25-14.96) |
| Ibrutinib | 4 | 58 (6) | 2 (50.0) | 2.08 (0.02-56.10) | 1 | 54 (0) | 1 (100.0) | na |
| **Chronic myeloid leukemia** | | | | | | | | |
| Tyrosine kinase inhibitors | 12 | 52 (10) | 5 (41.7) | 0.23 (0.14-0.50) | 10 | 52 (10) | 3 (30.0) | 0.17 (0.08-0.23) |
| **Acute myeloid leukemia and high-risk MDS** | | | | | | | | |
| Hypomethylating therapy | 5 | 59 (22) | 1 (20.0) | 0.04 (0.00-0.10) | 3 | 52 (29) | 1 (33.3) | 0.05 (0.00-0.07) |
| High-dose chemotherapy | 5 | 64 (2) | 3 (60.0) | 0.06 (0.01-0.24) | 4 | 64 (2) | 3 (75.0) | 0.18 (0.06-0.28) |
| **Myeloproliferative disease** | | | | | | | | |
| Ruxolitinib | 17 | 59 (8) | 9 (52.9) | 0.17 (0.05-0.59) | 12 | 58 (11) | 5 (41.6) | 0.11 (0.07-0.30) |
| **Allogeneic HCT** | | | | | | | | |
| < 6 months after HCT | 12 | 61 (11) | 5 (41.7) | 0.01 (0.00-0.28) | 9 | 65 (6) | 5 (55.5) | 0.19 (0.07-0.49) |
| Chronic GvHD | 13 | 61 (10) | 3 (23.1) | 0.11 (0.00-1.09) | 12 | 60 (10) | 3 (25.0) | 0.14 (0.00-0.81) |
| **CAR T cell therapy** | | | | | | | | |
| CD19-directed | 1 | 72 | 0 (0) | 0.00 (0.00-0.00) | 0 | na | na | na |

**Supplemental Table 1: Baseline characteristics of patients included in pseudovirus neutralisation analysis.** These were all patients with S1 IgG 50-300 BAU/ML and a random selection of patients with S1 IgG ≥300 BAU/ml after the standard 2-dose mRNA-1273 schedule (Haggenburg ea, Blood Advances 2022). [1]Absolute B cell number at time of inclusion; [2]absolute B cell number at time of 3rd vaccination. Na: not applicable.

### eTable 2

| | N | % | IgG S1 after 2nd vaccination (BAU/ml) median | 25th - 75th percentile | |
|---|---|---|---|---|---|
| **All patients** | 104 | 15.1 | 2323.0 | 529.1 | 5133.5 |
| **Lymphoma** | | | | | |
| During R-chemo | 3 | 7.0 | 0.1 | 0.1 | |
| <12 months after R-chemo | 3 | 7.7 | 10539.1 | 2812.7 | |
| <12 months after autologous HCT (BEAM) | 3 | 10.7 | 7012.2 | 68.1 | |
| **Multiple myeloma** | | | | | |
| 1st line therapy | 2 | 8.0 | 38.1 | 20.5 | |
| Daratumumab-containing therapy | 4 | 8.3 | 1310.3 | 291.2 | 2481.5 |
| IMiDs | 8 | 14.8 | 1129.0 | 357.3 | 16229.4 |
| <9 months after autologous HCT (HDM) | 4 | 8.3 | 3278.0 | 1942.2 | 3402.5 |
| **Chronic lymphocytic leukemia** | | | | | |
| Wait & see | 11 | 20.4 | 2934.1 | 307.4 | 9315.9 |
| Ibrutinib | 3 | 8.3 | 748.6 | 319.7 | |
| **Sickle cell disease** | | | | | |
| Hydrea containing therapy | 31 | 100.0 | 2920.8 | 1760.8 | 5567.8 |
| **Chronic Myeloid leukemia** | | | | | |
| Tyrosine kinase inhibitors | 10 | 19.2 | 2252.5 | 1447.5 | 3654.2 |
| **Acute myeloid leukemia and high-risk MDS** | | | | | |
| Hypomethylating therapy | 0 | 0.0 | | | |
| High-dose chemotherapie | 3 | 14.3 | 2842.2 | 1101.2 | |
| **Myeloproliferative disease** | | | | | |
| Ruxolitinib | 6 | 16.2 | 1909.4 | 614.2 | 5084.5 |
| **Allogenic SCT** | | | | | |
| < 6 months after HCT | 1 | 2.0 | 175.8 | 175.8 | 175.8 |
| Chronic GvHD | 5 | 8.9 | 8964.6 | 3149.3 | 17534.5 |
| **CAR T cell therapy** | | | | | |
| CD19-directed | 7 | 14.0 | 0.1 | 0.1 | 529.1 |

**Supplemental Table 2: Patients who did not receive a 3rd vaccination.** From the n=723 patients included in the cohort (Haggenburg ea, Blood Adv 2022), n=104 patients did not receive a 3rd vaccination. Only patients with hematologic malignancies were prioritized per Dutch COVID-19 vaccination protocol, and sickle cell disease patients (n=31) did not receive a 3rd vaccination within the timeframe of current analyses. The other patients (n=71) deferred the 3rd vaccination for personal reasons, most often because they felt they were sufficiently protected or they thought a 3rd vaccination would not be effective anyway. N: number of patients that did not receive a 3rd vaccination during the time under study.

**eTable 3**

| | SARS-CoV-2 infection | |
|---|---|---|
| | Baseline, n (%) | Follow up, n (%) |
| **All patients** | 24 (4.2) | 22 (2.0) |
| **Lymphoma** | | |
| During rituximab ± chemotherapy | 1 (2.6) | 1 (2.5) |
| <12mo after rituximab ± chemotherapy | 0 | 1 (2.8) |
| <12mo after autologous HCT (BEAM) | 0 | 0 |
| **Multiple myeloma** | | |
| 1$^{st}$ line therapy | 1 (4.3) | 2 (4.3) |
| Daratumumab | 1 (2.3) | 2 (2.3) |
| IMiDs | 5 (10.9) | 2 (2.2) |
| <9mo after autologous HCT (HDM) | 5 (11.4) | 1 (2.3) |
| **CLL** | | |
| Watch & wait | 0 | 3 (2.3) |
| Ibrutinib | 2 (6.1) | 1 (3.0) |
| **CML** | | |
| Tyrosine kinase inhibitor | 2 (4.8) | 4 (2.4) |
| **AML and high-risk MDS** | | |
| Hypomethylating therapy | 0 | 0 |
| High-dose chemotherapy | 0 | 0 |
| **Myeloproliferative disease** | | |
| Ruxolitinib | 2 (6.5) | 0 |
| **Allogeneic HCT** | | |
| < 6 mo after HCT | 1 (2.1) | 2 (2.0) |
| Chronic GvHD | 2 (4.3) | 2 (2.0) |
| **CAR T-cell therapy** | | |
| CD19-directed | 2 (4.8) | 1 (2.3) |

**Supplemental Table 3: SARS-CoV-2-infected patients.** Indicated are patients who had been SARS-CoV-2 infected prior to vaccination ('baseline') or became infected during follow up. Prior SARS-CoV-2 infection is defined as obtaining N IgG >14.3 BAU/ml at baseline or at any time during follow up. Mo: months.

### eTable 4

## A

| | All | IgG S1 after $2^{nd}$ vaccination (BAU/ml) | | | | Time (days)[1] | IgG S1 after $3^{rd}$ vaccination (BAU/ml) | | | | p-value |
|---|---|---|---|---|---|---|---|---|---|---|---|
| | N | N | median | 25th - 75th percentile | | Median (range) | N | median | 25th - 75th percentile | | $3^{rd}$ vs $2^{nd}$ |
| **All patients** | 27 | 25 | 443.1 | 31.4 | 3872.6 | 88 (1-140) | 26 | 540.8 | 60.0 | 8819.0 | 0.19 |
| **Autologous HCT** | | | | | | | | | | | |
| BEAM | 2 | 2 | 0.1 | 0.1 | | 33 (1-64) | 2 | 59.5 | 1.4 | | 0.18 |
| HDM | 14 | 13 | 562.6 | 130.8 | 3872.6 | 102 (30-140) | 13 | 8498.5 | 1315.4 | 12576.3 | 0.02 |
| Bu/Cy | 1 | 1 | 139.5 | 139.5 | 139.5 | 103 | 1 | 11767.1 | 11767.1 | 11767.1 | - |
| **Allogenic HCT**[2] | 7 | 6 | 832.4 | 144.0 | 6379.8 | 65 (41-106) | 7 | 73.5 | 49.0 | 109.2 | 0.05 |
| **CAR T cell therapy** | 3 | 3 | 0.1 | 0.1 | | 92 (43-119) | 3 | 0.3 | 0.1 | | 0.65 |

## B

| | N | % | Autologous HCT | Allogeneic HCT | CAR T cell |
|---|---|---|---|---|---|
| **All patients** | 27 | 4.6 | 17 | 7 | 3 |
| **Lymphoma** | | | | | |
| During R-chemo | 3 | 7.5 | 1 | - | 2 |
| <12 months after R-chemo | 1 | 2.8 | 1 | - | - |
| <12 months after autologous HCT (BEAM) | 1 | 4.0 | - | - | 1 |
| **Multiple myeloma** | | | | | |
| 1st line therapy | 14 | 60.9 | 14 | - | - |
| **Chronic lymphocytic leukemia** | | | | | |
| Ibrutinib | 1 | 3.0 | - | 1 | - |
| **Acute myeloid leukemia and high-risk MDS** | | | | | |
| Hypomethylating therapy | 3 | 18.8 | - | 3 | - |
| High-dose chemotherapie | 4 | 22.2 | 1 | 3 | - |

**Supplemental Table 4: Characteristics of patients who received cell therapy after the $2^{nd}$ mRNA-1273 vaccination. A.** Types of cell therapy received. [1]Time between cell therapy and $3^{rd}$ vaccination. [2]All allogeneic HCT recipients received lymphocyte replete transplants with a haplo-identical donor (n=3; 2 unvaccinated donors, 1 vaccinated), matched unrelated donor (n=3; vaccination status unknown) or cord blood donor (n=1). Six had GvHD prophylaxis (n=2) or GvHD therapy (n=4) at the time of $3^{rd}$ vaccination, which consisted of a calcineurin inhibitor or sirolimus, mycophenolic acid and/or prednisolone. BEAM: carmustine, etoposide, cytarabine, melphalan; HDM: high dose melphalan; Bu/Cy; busulphan cyclophosphamide. **B.** Cohorts of which intercurrent cell therapy patients were from.
