## supplemental figures for "Three-dose mRNA-1273 vaccination schedule: sufficient antibody response in majority of immunocompromised hematology patients"

### eFigure 1

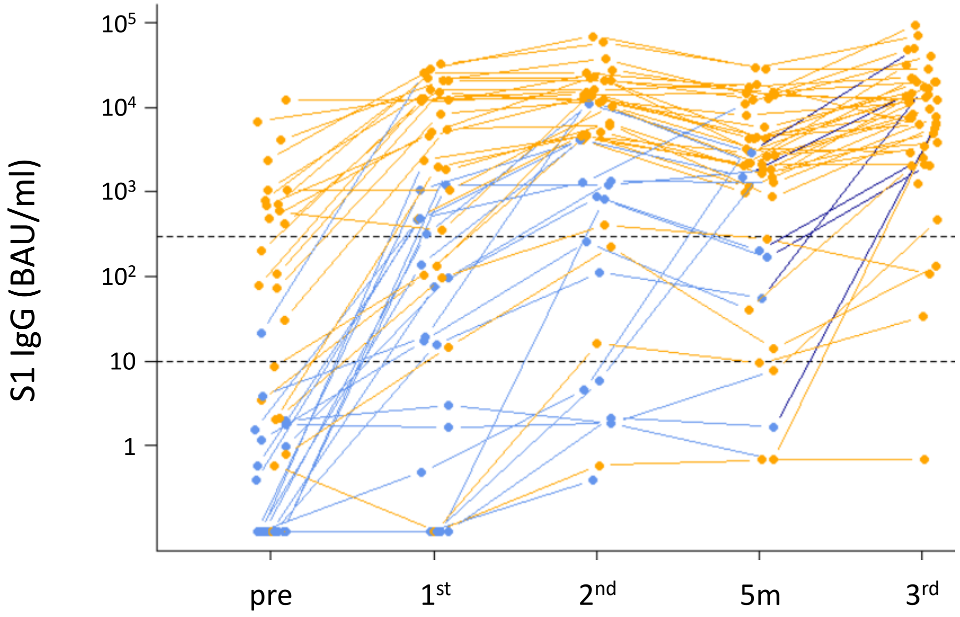


**Supplemental Figure 1. S1 IgG concentration in COVID-19 infected participants.** IgG S1 concentration at each timepoint for patients who were infected with SARS-COV-2 before the first vaccination or at any time during follow up. Dotted lines indicate seroconversion (S1 IgG >10 BAU/mL) and adequate S1 IgG concentration (≥300 BAU/ml). SARS-CoV-2 infection is defined as N IgG ≥14.3 BAU/mL at baseline or any time during follow up. Blue: N <14.3 BAU/ml at that timepoint; orange: N IgG ≥14.3 BAU/mL indicating SARS-CoV-2 infection at anytime before that timepoint.

**eFigure 2**

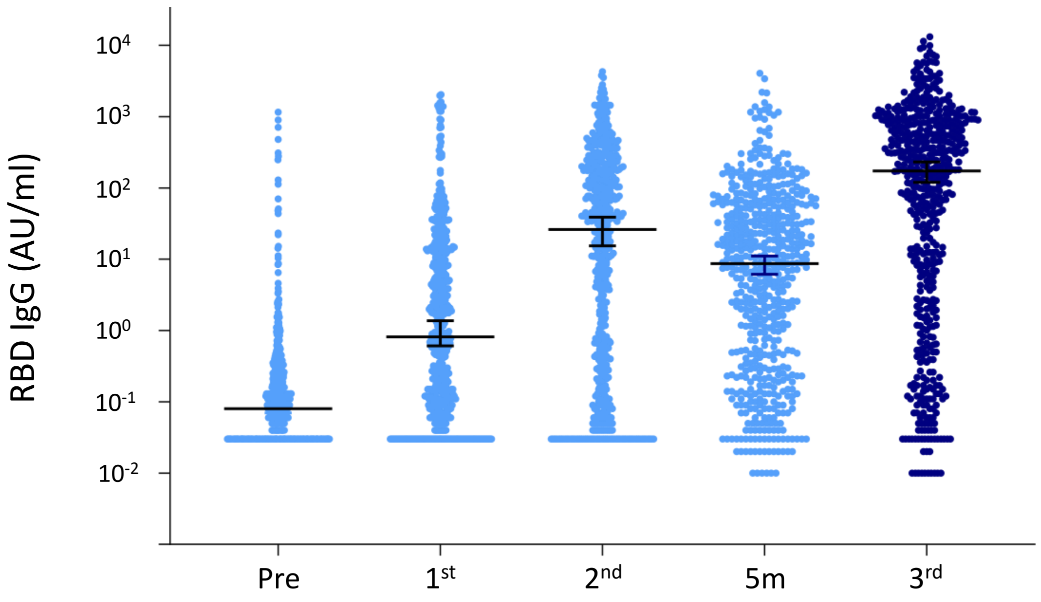


**Supplemental Figure 2. RBD binding antibody concentration.** IgG RBD concentration in AU/mL at each timepoint for previously uninfected patients (light and dark blue).
